# An Innovative Non-Pharmaceutical Intervention to Mitigate SARS-CoV02 Spread: Probability Sampling to Identify and Isolate Asymptomatic Cases

**DOI:** 10.1101/2020.10.07.20208686

**Authors:** Nathalie E. Williams, Xiaozheng Yao, Ankita Pal, Xiaolu Qian, Mansi Rathod, Chang Xu, Adrian Dobra

## Abstract

Studies estimate that a substantial proportion of SARS-CoV-2 transmission occurs through individuals who do not exhibit symptoms. Mitigation strategies test only those who are moderately to severely symptomatic, excluding the substantial portion of cases that are asymptomatic yet still infectious and likely responsible for a large proportion of the virus’ spread (*1-8*). While isolating asymptomatic cases will be necessary to effectively control viral spread, these cases are functionally invisible and there is no current method to identify them for isolation. To address this major omission in COVID-19 control, we develop a strategy, *Sampling-Testing-Quarantine* (*STQ*), for identifying and isolating individuals with asymptomatic SARS-CoV-2 in order to mitigate the epidemic. *STQ* uses probability sampling in the general population, regardless of symptoms, then isolates the individuals who test positive along with their household members who are high probability for asymptomatic infections. To test the potential efficacy of *STQ*, we use an agent-based model, designed to computationally simulate the epidemic in the Seattle with infection parameters, like R_0_ and asymptomatic fraction, derived from population data. Our results suggest that STQ can substantially slow and decrease the spread of COVID-19, even in the absence of school and work shutdowns. Results also recommend which sampling techniques, frequency of implementation, and population subject to isolation are most efficient in reducing spread with limited numbers of tests.

**Significance Statement:** A substantial portion of SARS-CoV-2 infections are spread through asymptomatic carriers. Until a vaccine is developed, research indicates an urgent need to identify these asymptomatic infections to control COVID-19, but there is currently no effective strategy to do so. In this study, we develop such a strategy, a procedure called S*ampling-Testing-Quarantine* (*STQ*), that combines techniques from survey methods for sampling from the general population and testing and isolation techniques from epidemiology. With computational simulations, we demonstrate that *STQ* procedures can dramatically decrease and slow COVID-19 spread, even in the absence of widespread work, school, and community lockdowns. We also find particular implementation strategies (including sampling techniques, frequencies of implementation, and people who are subject to isolation) are most efficient in mitigating spread.

## Introduction

With over six million confirmed cases, 190,000 deaths, and increasing prevalence in about 11 states at the date of this writing, the COVID-19 epidemic in the U.S. is far from controlled. This is despite the many symptomatic cases that have been identified, isolated, and treated and despite the near complete shut-down of the country, including businesses and schools, for months in Spring 2020. While we wait with bated breath for vaccines and treatments, the U.S., and indeed the world, is still in search of effective non-pharmaceutical interventions (NPIs). Without such NPIs, school districts and universities across the country are undertaking online classes in the fall and the calls for a return to shutdowns are increasing daily. All of these measures have shown to be partially effective, but at great cost to the economy and well-being of families. The key questions we seek to address in this study are: can less-disruptive non-pharmaceutical interventions substantially mitigate the COVID-19 epidemic and can they mitigate the epidemic as much as or more than disruptive interventions like school and work closures?

Instead of isolating the entire U.S. population for months on end, it would be much less financially, physically, and socially costly to isolate only those who are infected and only for the period in which they are infectious. The infectious cases that need to be isolated are comprised of symptomatic and asymptomatic infections. Individuals with symptomatic infections are already being tested, isolated, and treated when they present themselves for testing. It is the asymptomatic cases^1^, which are functionally invisible, that are confounding efforts at epidemic control. Indeed, studies suggest that as much as 79% of spread is through asymptomatic carriers and there could be anywhere from 3-40 times as many asymptomatic cases as symptomatic (*1-8*). Yet testing is largely reserved only for symptomatic cases. Scholars repeatedly warn that we are flying blind and new strategies are required to address this epidemic. Specifically, Li et al. find that “a radical increase in the identification and isolation of currently undocumented infections would be needed to fully control SARS-CoV2.” (*6*)

Current methods for identifying asymptomatic cases are universal and ongoing testing and thorough contact tracing. Universal and on-going testing is not efficient or feasible in the near term. Contact tracing has begun in some places, but multiple problems limit the efficacy of this strategy, including low contact rates, concerns about privacy and the speed of transmission (*9, 10*). In addition, given the substantial portion of transmission through asymptomatic or pre-symptomatic carriers, contact tracing can only be effective with identification of those infections which are currently functionally invisible.

To directly address these invisible asymptomatic infections, we propose and test the efficacy of an innovative procedure to identify and isolate individuals with asymptomatic infections. Our procedure, which we call *sampling-testing-quarantine* (*STQ*), combines the classic epidemiologic strategies of testing and quarantine with sampling strategies from survey methodology. The *STQ* procedure begins with probability sampling of the general population, regardless of symptoms. In this study, we use simple random sampling, cluster sampling, and pooled sampling strategies. Once sampled, individuals are tested for SARS-CoV-2 and those with positive tests are isolated, regardless of whether they exhibit symptoms or not.

We use an agent-based model (ABM) to test the efficacy of the *STQ* procedure for mitigating the spread of COVID in a computationally simulated population that mirrors that of Seattle Washington. While simplified portrayals of real populations, ABMs offer the ability to model the complex systems nature of human populations and provide the flexibility to experiment with various interventions on a simulated population in a way that would never be possible with real humans. Accordingly, ABMs are state-of-the art for testing the comparative efficacy of different intervention scenarios in epidemiology (*11-17*). They have been useful for understanding the spread of Ebola and Influenza (*17-20*) and to analyze the potential efficacy of various social distancing, lockdown, mask wearing and other non-pharmaceutical interventions on mitigating COVID-19 (*12-14, 16, 21, 22*). In particular, the ABM on which this study is based has been used to analyze the comparative efficacy of school closures and work from home policies on the COVID-19 epidemic in Seattle (*23*).

With this study, we examine multiple *STQ* intervention and non-intervention baseline scenarios. The baseline scenarios do not use *STQ* sampling of the general population and allow us to compare the difference in number of cases and dates of the highest number of cases between our *STQ* intervention scenarios and scenarios where no interventions are taken or where only symptomatic cases are isolated. Our *STQ* scenarios include multiple numbers of tests, and different frequencies at which the tests are applied. In this way, our results not only demonstrate the concept—that *STQ* interventions can have a substantial influence on mitigating the COVID-19 epidemic in the Seattle area—but also suggest which combinations of dates and tests might be most effective.

We also test a variety of sampling strategies to examine whether more sophisticated sampling strategies applied with the same number of tests can improve the efficiency with which *STQ* mitigates the epidemic. We use simple random sampling, where we randomly choose people from the population with equal probability. Our second method is cluster sampling, where we first separate people into the community clusters in which they live and separate children into the schools they attend and then randomly choose approximately the same proportion of people from each community or school cluster. Our third method is pooled sampling, where we place people in groups of two and groups of five, test a sample that is pooled from everyone in the group in the group, and isolate all people in the group if the pooled sample tests positive (i.e. if any one person in the group is infected).

For a comprehensive assessment, we also examine the mitigation potential of combinations of our *STQ* procedures (which isolate asymptomatic cases) with other less-disruptive non-pharmaceutical interventions. These include isolation of symptomatic cases and isolation of household members of positive cases.

## Results

Our results concentrate on the prevalence of symptomatic cases of COVID-19. While our focus in this article is to identify the asymptomatic cases that are integral to infection spread, it is the symptomatic cases that represent the morbidity and mortality outcomes that are better measures of epidemic severity.

### Demonstration of concept: can STQ mitigate the epidemic?

We begin with our most basic implementation of the STQ procedure, in order to analyze if it could be used to mitigate the severity and speed of COVID-19 spread. This is shown in Figure 1 which presents the prevalence and accumulation of symptomatic COVID cases for three baseline scenarios and three basic *STQ* intervention scenarios. Table S1 in the Supplementary Materials gives summary information of key outputs for these same six baseline and *STQ* scenarios.

**Figure 1.**
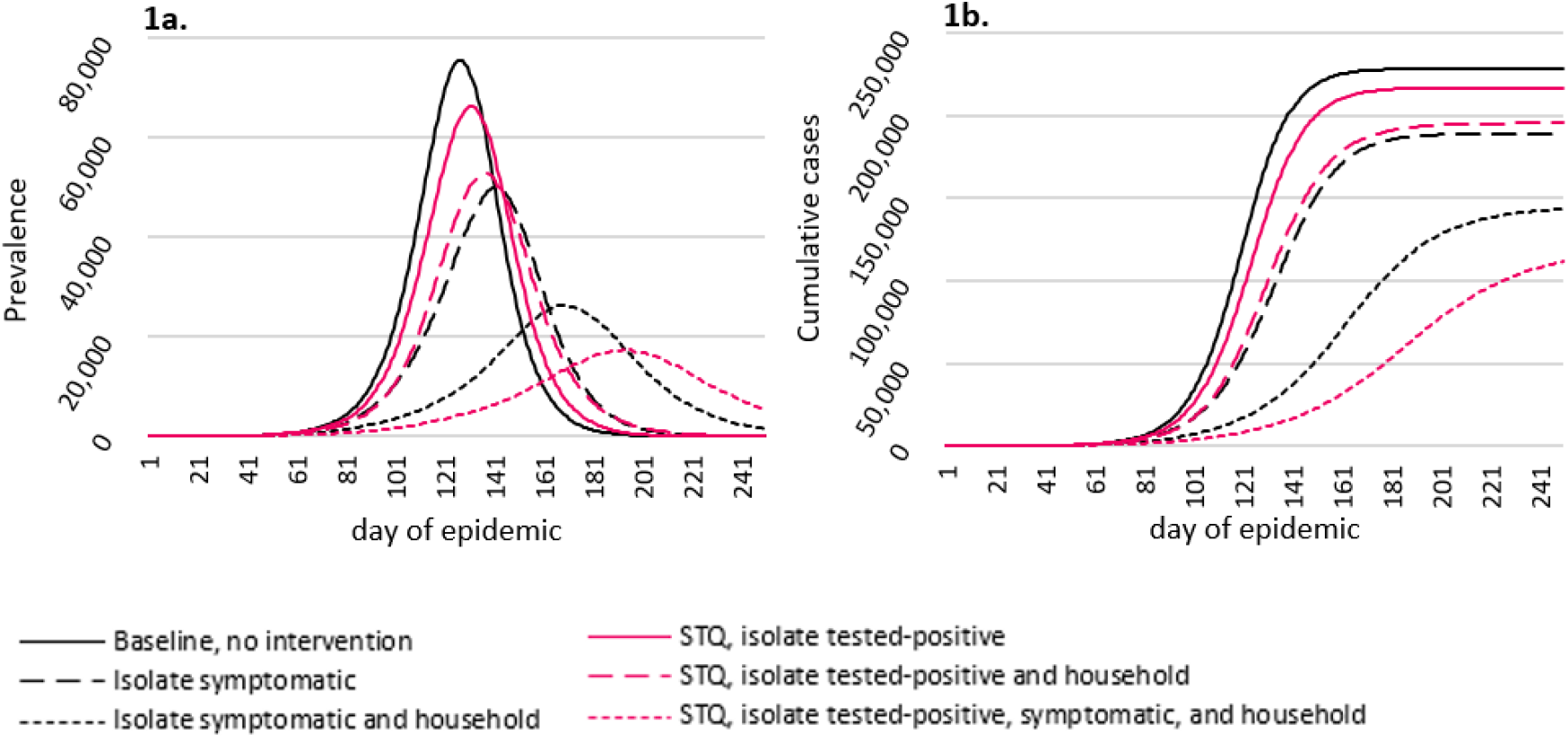
Efficacy of basic *STQ* interventions. Figure 1a shows prevalence of symptomatic infections. Figure 1b shows cumulative number of symptomatic infections. *STQ* scenarios used simple random sampling to select individuals for testing, 50,000 tests implemented once every seven days. Random seed used: 17392. See Table S1 for numerical presentation of these results.

As you can see, the peak height of the entirely unmitigated simulated epidemic (solid black line) reaches about 77,000 cases on day 136 of the epidemic. Isolating all symptomatic cases on the day they become symptomatic (dashed black line) reduces the peak to about 50,000 cases. This scenario notably reflects an ideal case of the status quo in the U.S. and many other countries, where people with symptoms are tested and asked to isolate. Isolating the household members of symptomatic individuals (dotted black line) further reduces peak prevalence to 26,000 cases. Already, it is clear that household members of symptomatic cases are an important pool of asymptomatic infections and isolating just those results in a substantial reduction in the epidemic.

The *STQ* procedure for identifying and isolating asymptomatic cases also substantially reduces the epidemic. Isolating only the asymptomatic cases who were sampled and tested positive reduces the peak prevalence to 66,000, isolating their family members to 53,000, and isolating the symptomatic and asymptomatic cases and their household members reduces the epidemic to about 18,000 cases at peak prevalence, just 24% of the peak of the unmitigated epidemic.

In addition, as visually shown in Figure 1a and numerically presented in Table S1, isolation of symptomatic, asymptomatic individuals, and their household members can delay the peak prevalence. As with numbers of cases, the largest delays in peak prevalence occur when household members are isolated along with symptomatic and known asymptomatic cases. When all known infected cases and their household members are isolated, this delays the peak prevalence by 74 days, almost two months, as well as decreasing the peak number of cases by 77% and the cumulative number of cases by 54%.

These results demonstrate the value of the *STQ* procedure: that random selection from the general population for testing (regardless of symptoms), can identify enough asymptomatic but infectious cases to reduce and delay the epidemic. Further, this sampling helps to identify household members of infectious cases who are also more likely than others to be infected.

### Improving the efficiency of STQ: number of tests, frequency of application, and sampling strategies

As shown in Figure 2, the number of tests used for the *STQ* procedure has a major impact on the prevalence of COVID-19. With just 10,000 tests used every seven days for the *STQ* procedure, the peak symptomatic prevalence decreases by two thirds, from approximately 77,000 to 25,000 cases, and is delayed by 50 days. With 100,000 *STQ* tests every seven days, peak prevalence further reduces to about 9,000 cases, and with 200,000 *STQ* tests to just 342 cases at the peak, which occurs later than 110 days later than the no intervention scenarios. Note that these results also include isolation of household members who are not tested as well as symptomatic agents. Our results clearly show that increased numbers of *STQ* tests and isolation of household members along with symptomatic and asymptomatic cases can decrease the peak symptomatic prevalence and cumulative symptomatic cases by over 99%. In other words, isolating only the infectious cases that can be found with our *STQ* procedure and the symptomatic cases that present themselves for testing, while allowing normal mobility for the remaining population, can dramatically reduce the COVID-19 epidemic.

**Figure 2.**
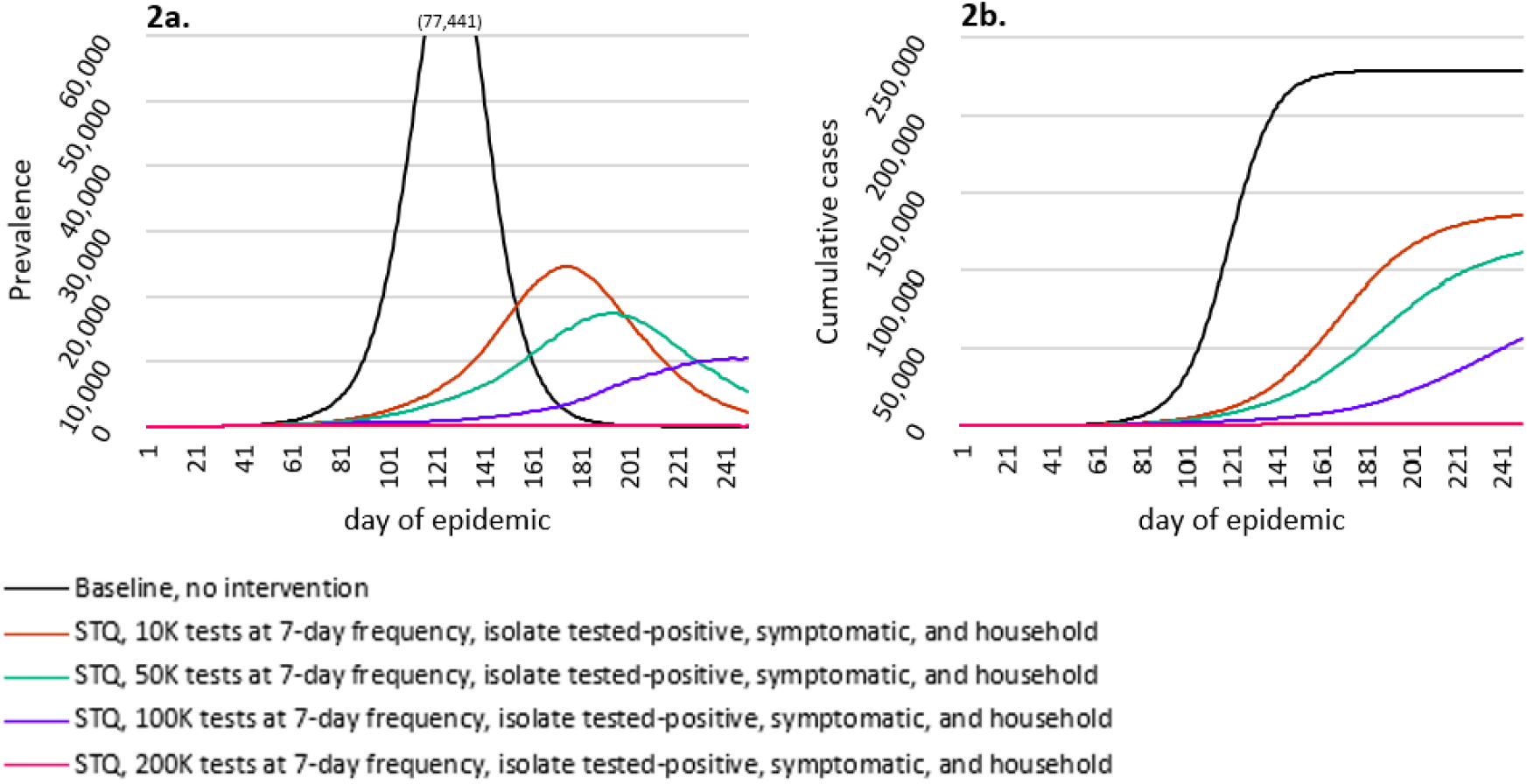
Efficacy of STQ intervention, by number of tests. Figure 2a shows prevalence of symptomatic infections. Figure 2b shows cumulative number of symptomatic infections. All *STQ* scenarios isolate the sampled-tested positive individuals, symptomatic individuals, and the household members of both; all used simple random sampling to select individuals for testing and tests were implemented once every seven days. Random seed used: 17392. The full simulated population of the Seattle ABM is 563,452. See Table S2 for numerical presentation of these results.

In Figure 3, we examine how best to employ the *STQ* procedure, by varying the number of tests and the frequency with which they are applied. For cases where a select number of tests are available, we address the question of whether it is best to use fewer tests more frequently or more tests less frequently. Our results suggest the best combination of number/frequency is larger numbers of tests employed at lower frequencies, which results in fewer symptomatic infections and a later date of peak symptomatic prevalence. This is likely because the initial implementation of all scenarios occurs on the same date (day 30 of the epidemic in this case) and larger numbers of tests employed at the initial date sets the epidemic onto a slower trajectory. However, it is notable that the outcomes of all four combination scenarios were relatively similar.

**Figure 3.**
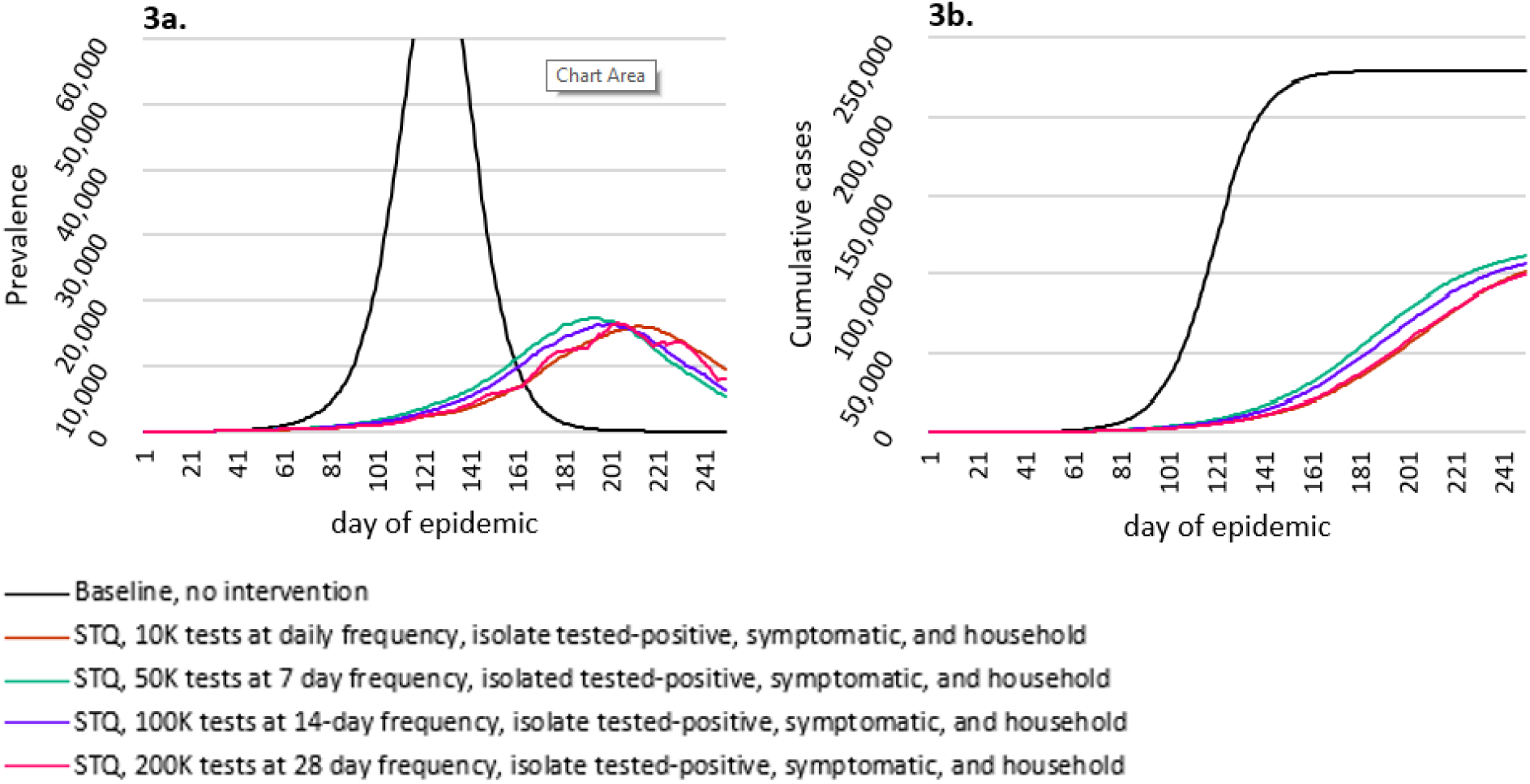
Efficacy of STQ interventions, by frequency of testing. Figure 3a shows prevalence of symptomatic infections. Figure 3b shows cumulative number of symptomatic infections. All *STQ* scenarios isolate the sampled-tested positive individuals, symptomatic individuals, and the household members of both; all used simple random sampling. Random seed used: 17392. See Table S3 for numerical presentation of these results.

Finally, we developed and tested the comparative efficacy of different sampling procedures for choosing the individual who would be tested. We present these results in Figure 4. As you can see, simple random sampling performs the worst, with higher peak symptomatic prevalence and cumulative symptomatic cases most iterations. Clustering on communities, where the same proportion of each community is selected for testing, performs about the same or even slightly worse when tested-positive and symptomatic agent and their household members were isolated. Under this full isolation scenario, the best sampling procedure where individuals are selected is clustering on schools, where the same proportion of each school is selected for testing and it is only children who are tested with *STQ* procedures. This likely occurs because of the lower rate of symptomatic cases in children; among adults there are a greater proportion of symptomatic cases and among children there are a greater proportion of asymptomatic cases. Thus, if we are searching for asymptomatic cases, it is best to search among children. Consequently, the combination of symptomatic cases, largely drawn from adults, and asymptomatic cases, entirely drawn from children and their household members through cluster sampling on schools, is the most effective individual-based intervention.

**Figure 4.**
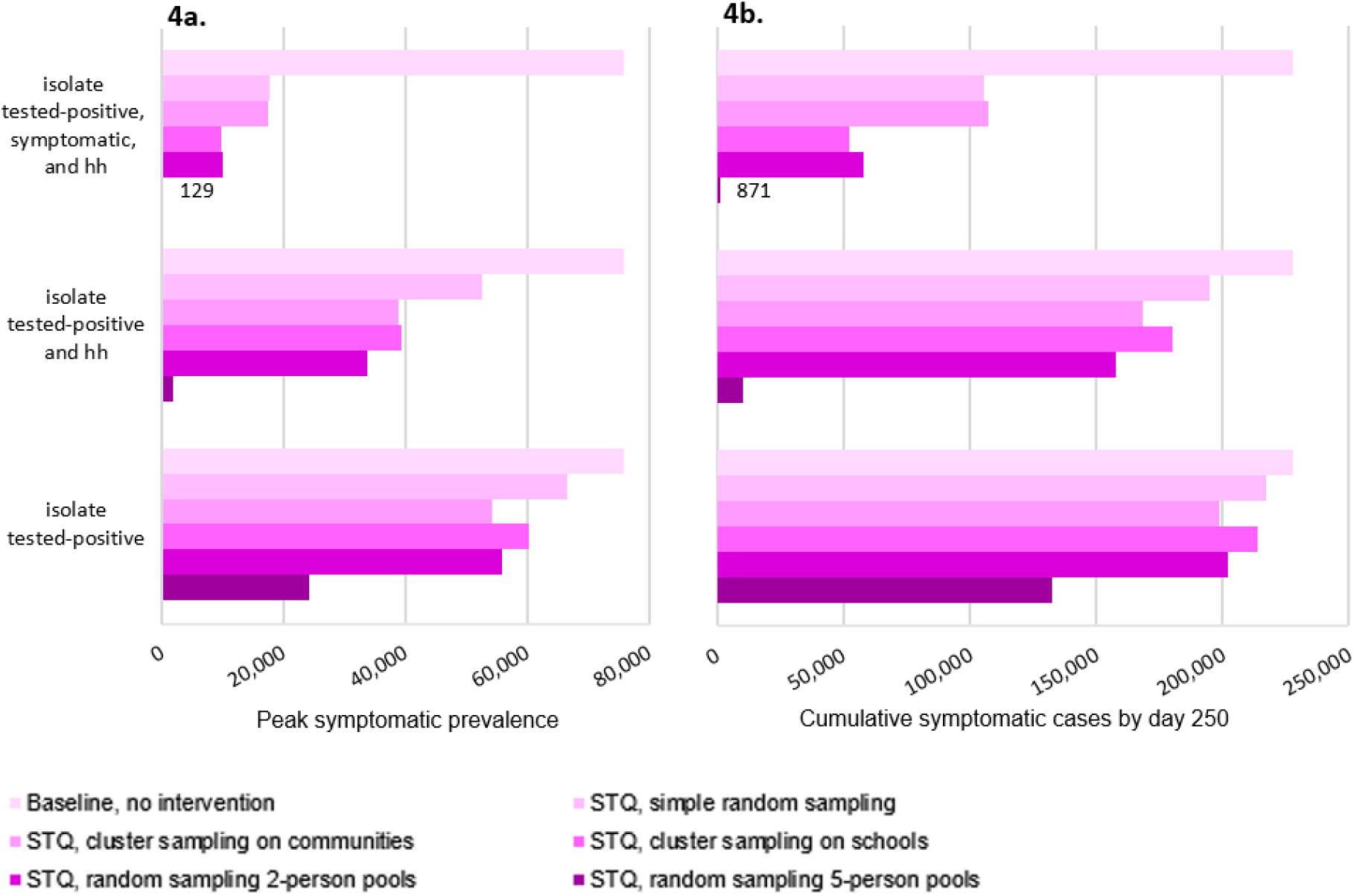
Efficacy of *STQ* interventions, by sampling strategy. Figure 4a shows highest daily prevalence of symptomatic infections. Figure 4b shows cumulative number of symptomatic infections at day 250. All *STQ* scenarios use 50,000 tests implemented once every seven days. Random seed used: 17392. See Table S4 for numerical presentation of these results.

Our pooled sampling scenarios are also likely effective. For both pooled sampling scenarios, we created groups of two people and five people. We used simple random sampling to select pools and each pool used a single test. If a pool tested positive (any one person in the pool was infected), then the entire pool was isolated, along with their household members. In this way, we use the weakest sampling strategy (simple random sampling), but reach two and five times as many people. Pooled sampling with 2-person pools had similar efficacy to individual sampling that was clustered on schools. However, pooled sampling with 5-person pools is the most effective strategy we find, with peak symptomatic prevalence at 12, a cumulative total of only 871 cases by day 250.

The model results we present here are ideal case scenarios. These are intended to demonstrate the concept, that 1) identifying and isolating asymptomatic cases can substantially mitigate the COVID-19 epidemic, and 2) that sampling in the general population for testing and isolating untested household members are methods to accomplish this. Our ideal scenarios assume that 100% of sampled agents agree to be tested, and that all sampled and tested-positive agents will quarantine with complete efficacy. It also assumes that all symptomatic agents will get tested and quarantine with complete efficacy, which we already know is not the case in the U.S. population.

If implemented in the U.S. or elsewhere, we can assume that the efficacy of the *STQ* procedure will be less than estimated here, due to lower than 100% response rates, testing inaccuracies, and incomplete isolation. Indeed, response rates to surveys in the U.S. vary dramatically with the best rates achieving about 90% response. The lower efficacy of the *STQ* procedure under these conditions can be simply estimated from our results at hand. For example, if 100,000 tests are used, but the response-quarantine-efficacy rate is 50%, then the estimated mitigation of this scenario will be the same as the scenario where 50,000 tests are used at a 100% response/quarantine-efficacy rate. In this scenario, we still find substantial mitigation of the COVID-19 epidemic.

### Efficiency

In summarizing the above results, the best *STQ* strategy for mitigating the greatest number of symptomatic cases and deaths is to use the largest number of tests (to cover the full population) on a daily basis. Given that this is not currently feasible, for policy relevance we instead calculate the cost effectiveness of various *STQ* strategies in terms of the number of tests required to avert one symptomatic case. As shown in Table 1, we use CDC data^2^ to estimate that one out of every 18 tests result in identification of one positive COVID-19 case. From our model results, we then estimate that if the household members of symptomatic cases are also isolated (without testing them), a much lower eight tests are required to avert each one symptomatic case. Moving to the *STQ* scenarios, using simple random sampling and isolating only the cases that test positive with STQ results in 145 tests required to avert one symptomatic case. This decreases to a low of 16 tests to avert one symptomatic case for pooled sampling of 5-person pools and seven tests for pooled sampling of 5-person pools if household members are also isolated. Notably, the *STQ* scenario of pooled sampling of 5-person pools is slightly more efficient than the current status quo of testing and isolating symptomatic cases. All other *STQ* scenarios are less efficient than the status quo. However, instituting even these less efficient *STQ* scenarios is likely to avert a substantial number of cases (as described above) and could be more cost effective than the emergency room visits, long-term care, lost labor, and other economic costs of symptomatic cases and deaths. These comparisons are intended to demonstrate the possible efficiency of *STQ* procedures and not to imply that STQ should entirely replace testing of symptomatic individuals.

**Table 1.** Estimated number of tests to avert one symptomatic case.

|  | Isolate infected case only | Isolate infected case and all household members |
| --- | --- | --- |
| Isolate symptomatic * | 18 | 8 |
| STQ- Simple random sampling | 145 | 48 |
| STQ- Cluster sampling on communities | 82 | 29 |
| STQ- Cluster sampling on schools | 113 | 34 |
| STQ- Pooled sampling, 2-person pools | 62 | 23 |
| STQ- Pooled sampling, 5-person pools | 17 | 7 |

\* Calculations for isolate symptomatic based on CDC estimates of positive test rates. All other calculations based on model results. Details on calculations available in Supplementary Online Materials.
<sup>1</sup> All scenarios herein use 50,000 tests, once every seven days.

As shown in Table 1, isolating household members of infected cases is much more efficient than not. We calculated the exact multiplicative effect of household member isolation for each of our scenarios. As shown in Table 2, isolating household members of individuals who experience symptoms is estimating to avert 2.22 times more symptomatic cases than not isolating them. The multiplicative effect is slightly higher for *STQ* scenarios and highest for cluster sampling on schools, where 3.37 times more symptomatic cases are averted by isolating household members.

**Table 2.**
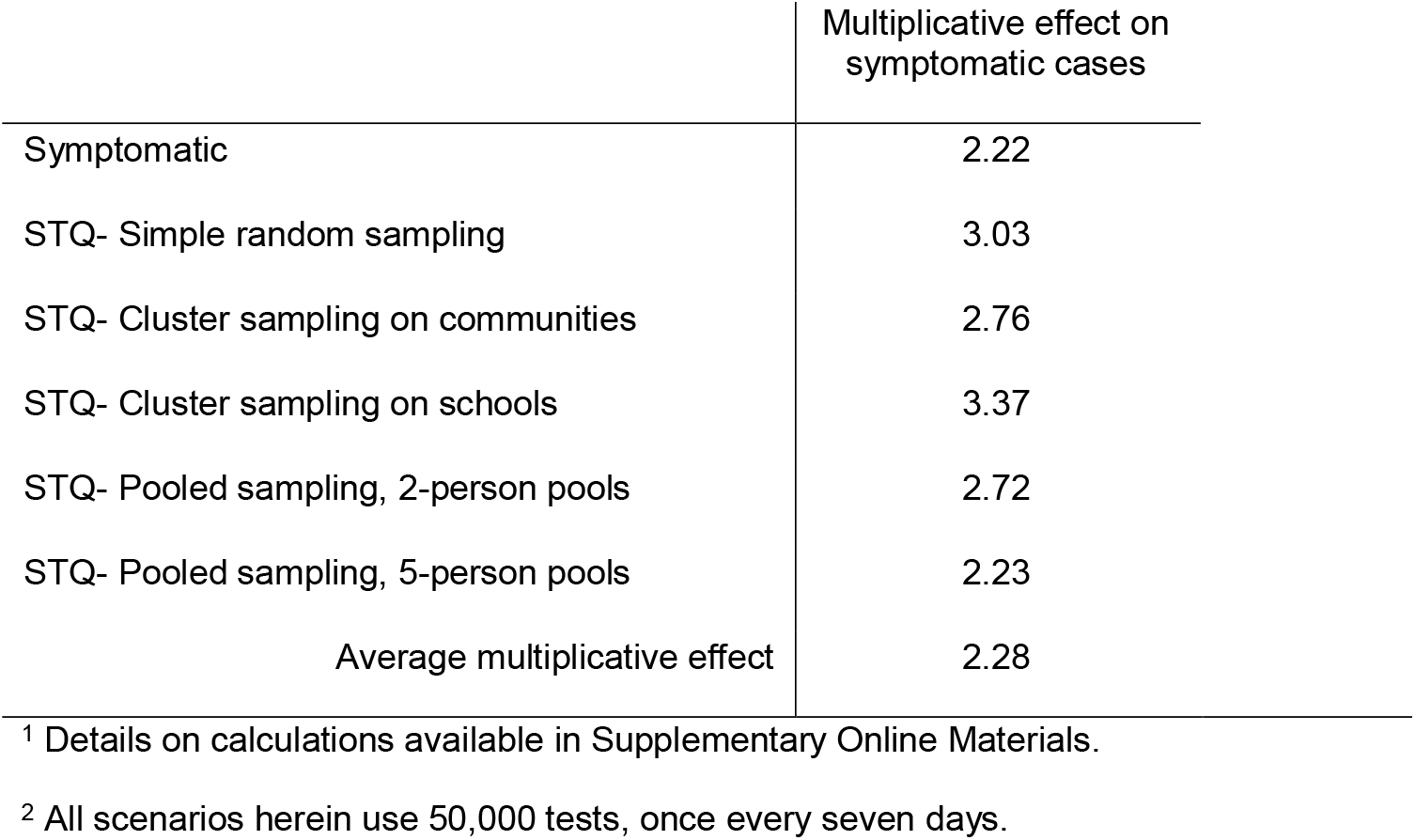
Multiplicative effect of isolating household members

## Discussion

The results we present here suggest that our *STQ* procedure has the potential to substantially reduce and delay the spread of the COVID-19 epidemic, as well as reduce the morbidity and mortality tolls. This could be accomplished without the disruptions of school, work, and complete shutdowns or it could be combined with other non-pharmaceutical interventions (like masks and shutdowns) to create even more effective mitigation. The key conclusions from this study are that sampling from the general population can be effective for identification and isolation of asymptomatic cases and that isolating household members of known infected individuals is an efficient and effective method for further slowing spread through asymptomatic cases. In addition, our results suggest strategies to improve the efficiency of the *STQ* procedure for greater mitigation of the epidemic with fewer tests. These include cluster sampling, sampling among the child and youth population that has a greater proportion of asymptomatic cases, and sampling pools of people instead of individuals. Our goal with this study is to demonstrate the possible efficacy of general population sampling, with techniques derived from survey methodology, and initiate the search for even more efficient methods to identify the asymptomatic cases that are integral to the spread of COVID-19. Those asymptomatic cases have largely been ignored in current strategies for epidemic mitigation and, with our results in hand, it is time to change such omissions.

That said, none of our model outcomes that are shown in this paper should be taken to imply that the scenarios we test here will result in the exact number of symptomatic infections in the real population of Seattle, or the exact percentage changes in prevalence in any other city. Our ABM, as with all computational, mathematical, and statistical models, is a simplified portrayal of the complexity of real human populations. Instead, given the sensitivity testing we undertook with a variety of parameters for the infectiousness and virology of SARS-CoV-2, our results suggest the general direction of difference between two intervention scenarios, or the comparative advantage of one intervention over another. Thus, we do not expect that the scenario that uses simple random sampling, 100,000 tests, employed every 14 days and isolates symptomatic cases and sampled/positive cases and their household members will result in exactly 131,866 fewer symptomatic cases than the no intervention scenario. We do expect that this scenario will result in substantially fewer cases than if no interventions are applied and we also expect that it will result in fewer cases than the scenario that applies 50,000 tests every seven days.

We specifically focus on the case of Seattle Washington here as a discrete and relatively homogenous population (being entirely urban with high density and urban-reflective mobility patterns) with which to undertake this demonstration of the *STQ* concept. However, our results can also be reasonably interpreted to represent the possible mitigation potential of the *STQ* procedure in other urban areas with similar density and mobility patterns. With the demonstration of concept verified, we also argue that *STQ* will likely be effective in other suburban and rural areas or on a country level, but the exact magnitude and speed of the mitigation and the most effective sampling strategies might be different in those cases. Further modeling will be required to estimate such outcomes.

Even with our results that demonstrate efficacy of the *STQ* procedure, it could be reasonably argued that randomly choosing people from the population to be tested and then isolating them, their untested family members, and the possibly uninfected people in a pooled sample is a draconian policy. At the same time, the COVID-19 epidemic has laid bare the altruistic potential of many people around the globe who have already been willing to isolate themselves, to great discomfort and difficulty, for the sake of others, epitomized by the now trite statement “we’re all in this together”. Thus, we intend for our modeled scenarios to be taken as an opportunity to allow for voluntary participation in a likely effective program that could substantially decrease the spread, prevalence, and deaths from COVID-19. In addition, our ABM models an isolation period of two weeks, which is almost a magnitude of 10 shorter than the social distancing and quarantine that many Americans have already voluntarily instituted for themselves. Even further, a full implementation of the *STQ* procedure could include substantial support for those participants that are tested and voluntarily isolate themselves and household members, including food and goods delivery, nursing and medical care, academic support for children, and paid sick leave. The cost of such extensive support would still likely be less than the cost of the current and continuing quarantine on the U.S. economy.

While the cost of the *STQ* suggested quarantine could be mitigated for individuals and their families, we are also aware that an *STQ* program could be expensive. It would include the cost of hundreds of thousands of test kits, sampling statisticians, and nurses/enumerators to undertake the tests. However, the initial cost of the partially mitigated COVID-19 epidemic is already in the trillions of dollars in the U.S. alone and the economic impact of historic highs in unemployment has only begun to be understood. In comparison, the cost of a *STQ* intervention program could be calculated with reasonable simplicity and would in all likelihood have a much smaller long-term price-tag than no such intervention.

In addition to mitigating the spread of COVID-19 and the economic costs, another benefit of the *STQ* procedure would be much more accurate estimates of the epidemiology of the SARS-CoV-2 virus. One of the difficulties of current estimates of the infectiousness, symptomatic fraction, prevalence, and death rates related to SARS-COV-2, as well as inequalities in all the above, is that they are based on data that largely include only the moderately to severely symptomatic people who are tested. In other words, current estimates are based on biased samples. Thus, even with the best statistical and mathematical minds performing incredible numerical gymnastics, the result is large confidence intervals and wide ranges of possible values for each estimate. Alternately, the *STQ* procedure, based on probability sampling techniques applied to the general population (which includes people who are symptomatic, asymptomatic, and not infected), would provide the best possible unbiased data for calculating much improved estimates for COVID-19 spread.

Throughout the COVID-19 epidemic, there have been concerns that more testing will uncover more infections and make the epidemic look worse than if fewer tests are used. However, we show exactly the opposite. The *STQ* procedure that we developed and tested in this study requires many more diagnostic tests than are currently being instituted. At the same time, *STQ* provides a clear procedure for turning more tests into better numbers. Our results suggest that when implemented with proper sampling techniques, at particular intervals, and paired with two-week quarantines at the individual or household level, the more testing that is done, the lower the case numbers will look and the lower they actually will be.

## Materials and Methods

### CORVID Agent-Based Model

The agent-based model (ABM) upon which are study is based, CORVID, was developed by Dennis Chao, available at https://github.com/dlchao/corvid and described in detail at (*23*). The model simulates the population of Seattle, WA, with data on population residences, employment, schooling, and mobility from the year 2000. The CORVID population of agents are separated into communities, of about 2000 people each. School-aged agents attend schools with other agents in their communities. Adult-aged agents work at employers with agents from their own and other communities. Agents also interact with others within their communities.

There are two daily time steps: the day-time step where individuals interact with and can be infected by agents within their school, employer, or community and the night-time step where agents interact with and can be infected by others within their household. Infections occur when two individuals interact. The probability that an interaction with an infected agent results in transmission of the infection is governed by β, which is 0.168206 for models with R_0_=2.6, 0.127852 for models with R_0_=2.0, and 0.22201 for models with R_0_=3.4. Results in this paper reflect R_0_=2.6, a commonly used value at this time, and results for a broader range of possible values, R_0_=2.0 and 3.4 (*2, 24-27*), are presented in the Supplementary Materials.

### STQ Simulations

We tested six series of *STQ* interventions. Series A includes no sampling. Scenario A1 has no interventions, a completely unmitigated epidemic. A2 isolates all agents who are symptomatic on the day they develop symptoms. This is an ideal case of the current status quo in the United States and many other countries, where only individuals with symptoms are tested and isolated. A3 isolates symptomatic agents and their household members. All isolations are completely effective (no chance of transmission to any other agent) and last until the agent is no longer infectious (two weeks in the CORVID model).

The B series of scenarios uses simple random sampling in the general population, regardless of symptoms, to select individual agents for testing. If a selected agent tests positive, they are isolated with complete efficacy for two weeks. The C series uses cluster sampling on communities. The same proportion of each community is selected for testing. Once the community proportion is calculated, individual agents from each community are randomly selected for testing. The D series uses cluster sampling on schools. The same proportion of each school is selected for testing, except in cases where schools are small, in which case at least one student is selected from each school. Because the D series concentrates on schools, only school-aged agents are selected for testing and adults are never selected. The E series uses pooled sampling with pools of two people each. All agents are placed into pools of two people with others in their community. Pools are randomly sampled from the population for testing. If a pool tests positive (i.e. either agent in the pool is infected), then both agents in the pool are subject to isolation. The F series uses pooled sampling with pools of five people each. All agents are placed into pools of five people with others in their community. Pools are randomly sampled from the population for testing. If a pool tests positive (i.e. any one one agent in the pool is infected), then all agents in the pool are subject to isolation.

For all series B-F, we ran three scenarios. The first (B1, C1, etc.) isolates only the agents who are sampled with *STQ* procedures and test positive (“tested-positive” agents). The second (B2, C2, etc.) isolates the tested-positive agents and their household members. The third (B3, C3, etc.) isolates the tested-positive agents, symptomatic agents, and the household members of both. Details and code for the *STQ* scenarios are available at: https://github.com/CORVID-ABM

## Data Availability

Model code and all related data are available on github, as noted in the manuscript.

https://github.com/CORVID-ABM

## Acknowledgments

The authors greatly appreciate Dr. Dennis Chao, who created the original *CORVID* agent-based model that we use for this study. We also thank the Department of Sociology at the University of Washington for funding to support programming efforts for this study.

## Supplementary Information for

### Supplementary Information Text

#### Calculations for Table 5

Table 5 presents a measure of efficiency of each scenario, specifically the estimated number of tests necessary to avert one symptomatic case of SARS-CoV-2. For the non-*STQ* scenario where only symptomatic cases are isolated, an ideal case of the current status quo in the U.S., we calculated this using the CDC reported percent of respiratory specimens that test positive for SARS-Cov-2, which is equal to 5.5 at the most recent reporting^3^. If 5.5% of tests (largely taken from symptomatic individuals) are positive, then about one out of every 18 tests is positive. For the non-*STQ* scenario where household members are also isolated, we divided the 18 tests by the multiplicative effect of isolating household members on the number of symptomatic cases averted that we calculated from our ABM simulations as the number of cases averted when household members are isolated, divided by the number of cases averted when household members are not isolated.

For all *STQ* scenarios, the tests required per symptomatic case averted is calculated with the total number of tests used (e.g. 50,000 tests every seven days, starting on day 30 and ending day 250 = 1,600,000 tests), divided by the number of symptomatic cases averted. The number of symptomatic cases averted is calculated by subtracting the cumulative symptomatic cases at day 250 in the STQ scenario from that in the baseline/no intervention scenario.

#### Calculations for Table 6

Table 6 presents the multiplicative effect of isolating household members of infected cases. We calculated this by dividing the number of symptomatic cases averted in the scenario that isolates infected cases and household members by the number of symptomatic cases averted in the scenario that isolates only infected cases. The number of symptomatic cases averted is calculated by subtracting the cumulative symptomatic cases at day 250 in the STQ scenario from that in the baseline/no intervention scenario.

#### Tabular results to support Figures 1-4 in the main manuscript

The following tables present numerical results from the same simulations that are shown in Figures 1-4 in the main manuscript. These tables include additional information, such as the reductions in the number and percent of cases from the non-intervention scenario and the dates of peak prevalence.

#### Multiple seed simulations and presentation of results

For each scenario, we ran 10 simulations. Each simulation was initiated with a different random seed, to allow for stochasticity in results even within the same scenario. In all scenarios, outputs from simulations with different random seed starts result in the same substantive conclusions, namely that: the STQ procedure can be an effective non-pharmaceutical method to substantially mitigate the COVID-19 epidemic; cluster sampling on schools and pooled sampling are more efficient than simple random sampling; a larger number of tests at less frequent intervals are the most efficient; and isolating household members, symptomatic agents, and asymptomatic agents is the most efficient strategy.

As shown in Figure S1, there was between scenarios with different random seed starting points. There was very slight variation in the peak prevalence and in the shape of prevalence curves for scenarios where the frequency of testing was two and four weeks. The greater variation was in the date of peak prevalence.

#### Variations in R_0_

There are still many uncertainties in the scientific community around key parameters about SARS-CoV-2 spread, including our understanding of R_0_. Studies estimate a wide range of R_0_, depending on the characteristics and location of the study population (*1-3*). The results presented in our main text use an R_0_ of 2.6. For a full range of possible effects, we also tested all simulations with a low R_0_ = 2.0 and a high R_0_ = 3.4, a range similar to several other studies and as recommended by the CDC (*2, 4, 5*). As shown in Figures S2 and S3, *STQ* interventions can substantially mitigate the epidemic for all values R_0_. The percent of infections mitigated is greater for lower values of R_0_.

**Fig. S1.**
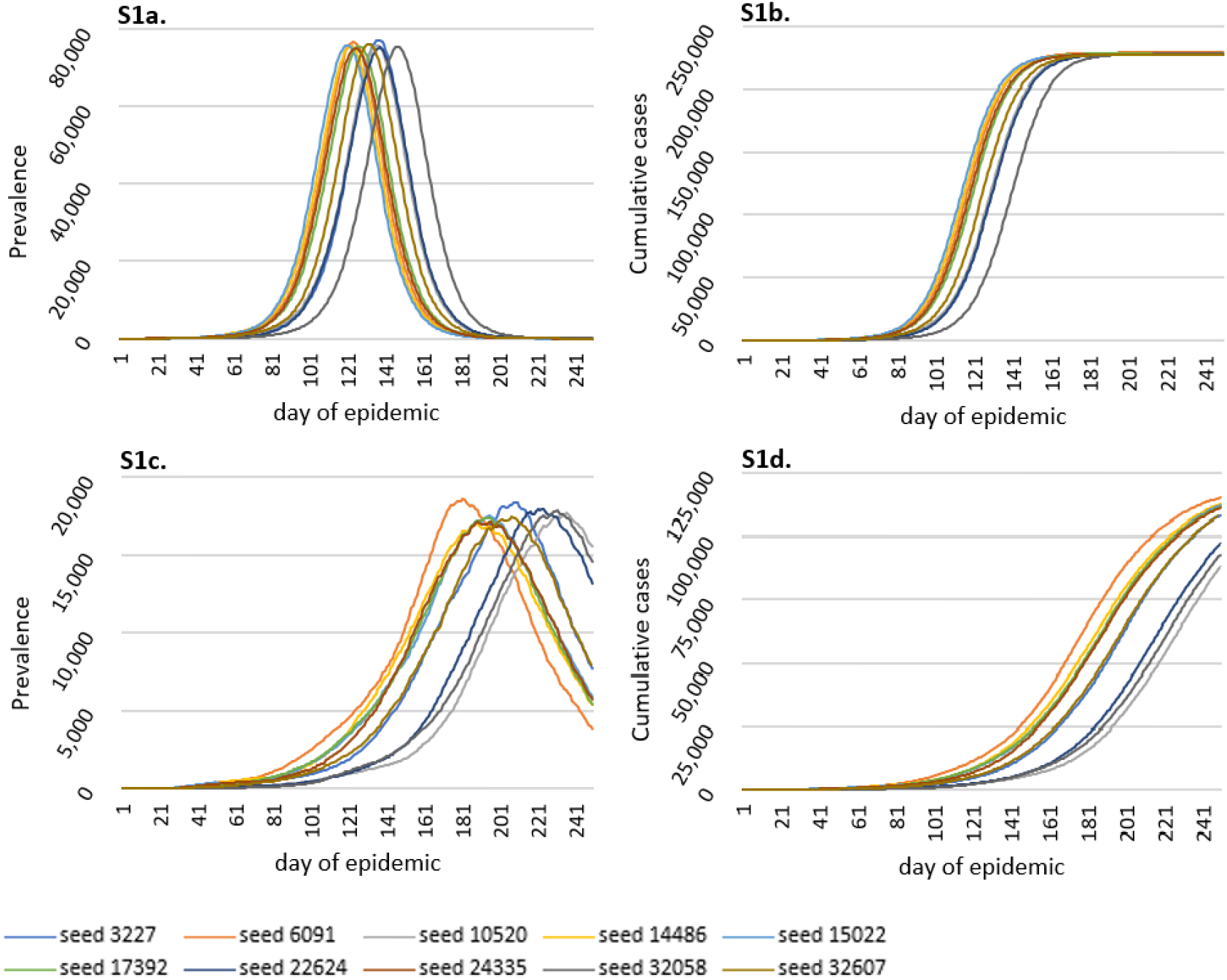
Variation between scenarios when different random seeds are used. Figures S1a-b scenarios are the baseline with no interventions, with S1a presenting symptomatic prevalence and S1b cumulative symptomatic cases by day 250. Figures S1c-d show *STQ* scenarios with 50,000 tests implemented every seven days, simple random sampling, and isolation of tested-positive, symptomatic and household members of both. S1c presents symptomatic prevalence and S1d cumulative symptomatic infections by day 250.

**Fig. S2.**
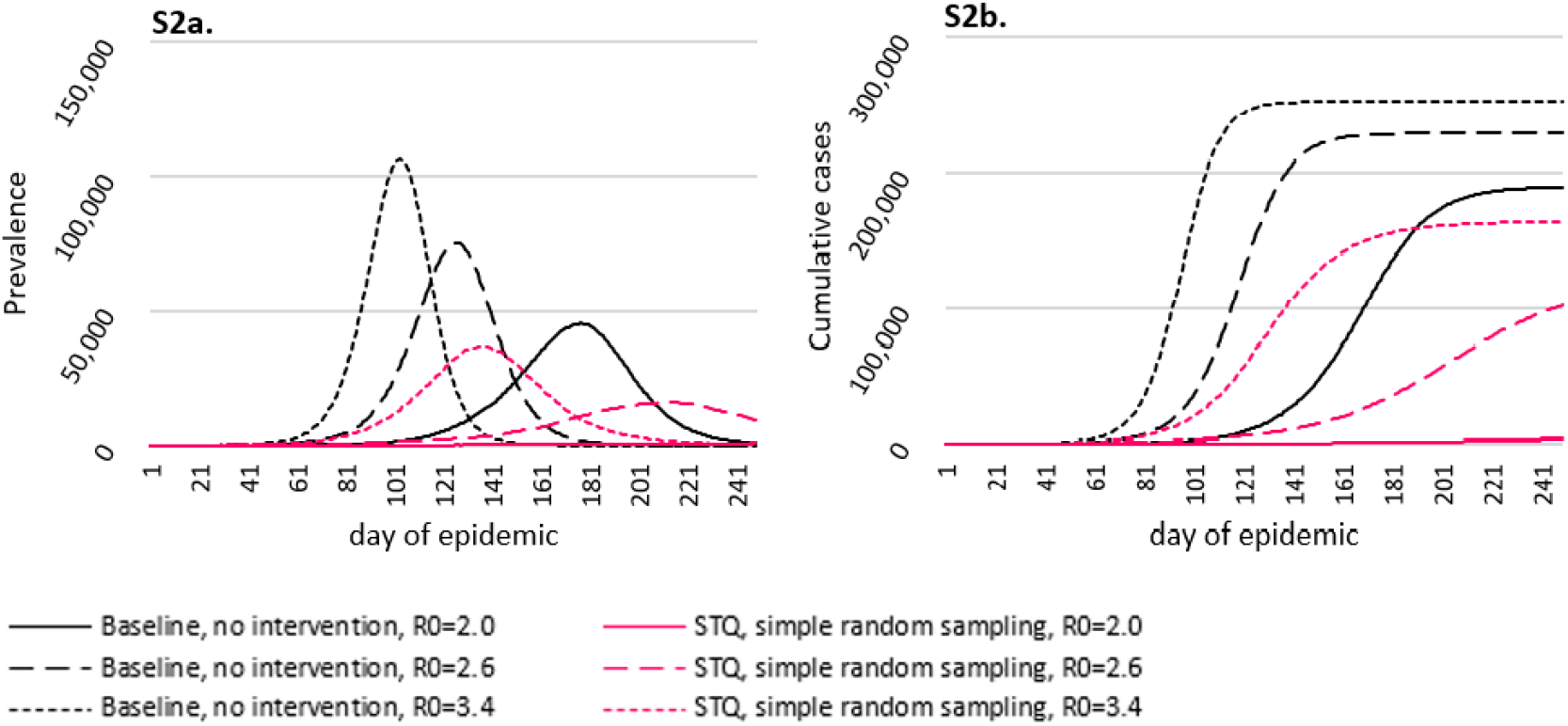
Variation between scenarios when different R_0_ values are used. Figure S2a shows symptomatic prevalence scenarios and S2b cumulative symptomatic cases by day 250. Black lines represent the baseline, no intervention scenario. Red lines represent an STQ scenario with simple random sampling, 50,000 tests implemented every seven days, isolation of tested-positive, symptomatic, and household members of both, and seed 17392.

**Fig. S3.**
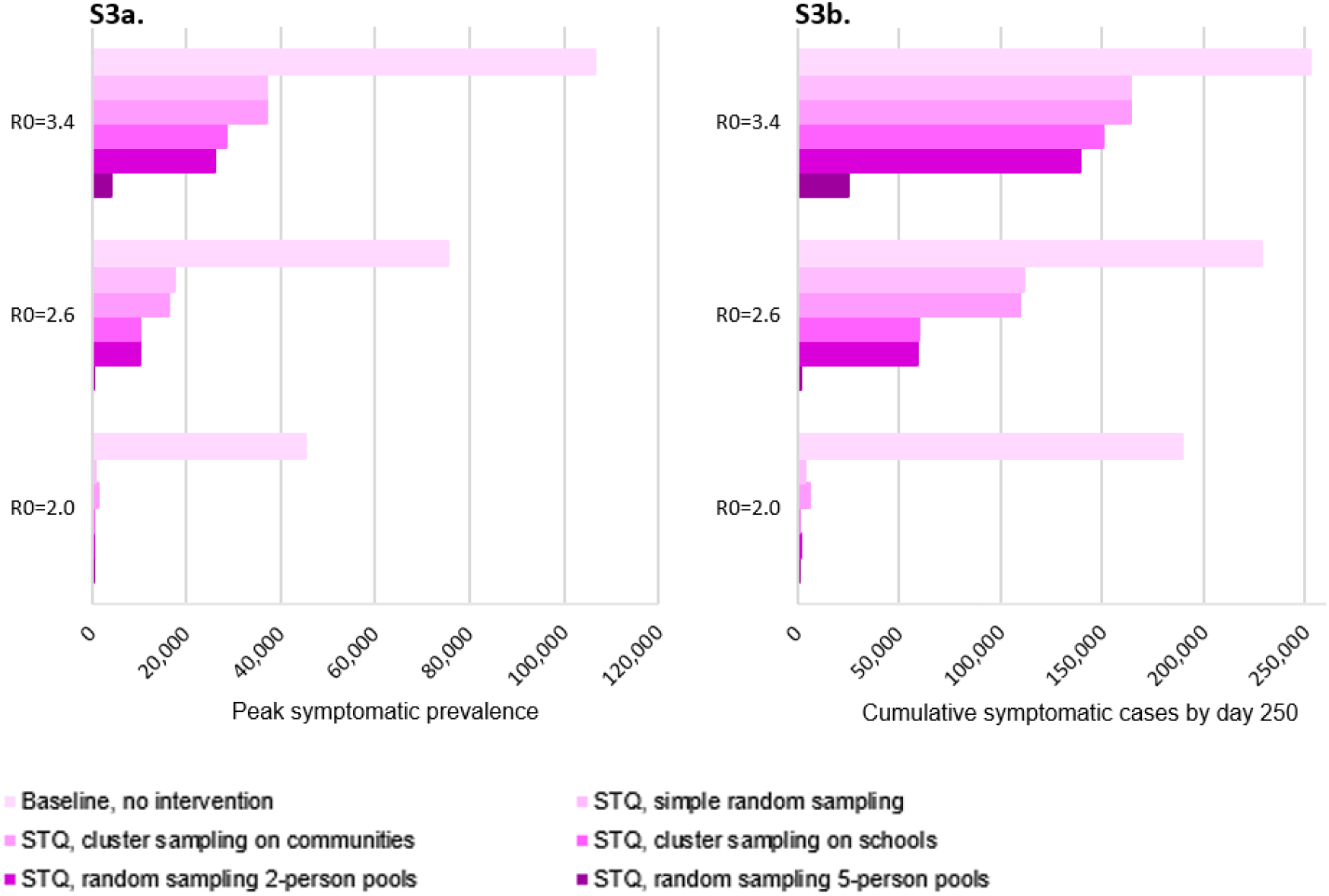
Variation between different sampling strategies when different when different R_0_ values are used. Figure S3a shows peak symptomatic prevalence and S3b cumulative symptomatic cases by day 250. All *STQ* scenarios use 50,000 tests implemented every seven days, isolation of tested-positive, symptomatic, and household members of both, and seed 17392.

**Table S1.** Efficacy of basic STQ interventions. Scenarios here are the same as presented in Figure 1 in the main article.

|  | Symptomatic prevalence at peak |  | Cumulative symptomatic cases at day 250 |  | Date of peak symptomatic prevalence |  |
| --- | --- | --- | --- | --- | --- | --- |
|  | (# cases) | Reduction from baseline<br>(#/% cases) | (# cases) | Reduction from baseline<br>(#/% cases) | (Date of epidemic) | Delay from baseline<br>(# days) |
| Baseline, no intervention | 75,809 |  | 228,030 |  | 131 |  |
| Isolate symptomatic | 49,770 | 26,039 / 34% | 189,334 | 38,696 / 17% | 149 | 18 |
| Isolate symptomatic, and household | 26,488 | 49,322 / 65% | 142,194 | 85,836 / 38% | 177 | 45 |
| STQ, isolate tested-positive | 66,563 | 9,246 / 12% | 216,980 | 11,050 / 5% | 138 | 7 |
| STQ, isolate tested-positive and household | 52,487 | 23,322 / 31% | 194,622 | 33,408 / 15% | 147 | 16 |
| STQ, isolate tested-positive, symptomatic, and household | 17,683 | 58,126 / 77% | 105,643 | 122,387 / 54% | 205 | 74 |

<sup>1</sup> All values represent the average of 10 model runs, each starting with a different random seed.
<sup>2</sup> All STQ scenarios used simple random sampling, 50,000 tests, every seven days.

**Table S2.** Efficacy of STQ interventions, by number of tests used. Scenarios here are the same as those presented in Figure 2 in the main article.

|  | Symptomatic prevalence at peak |  | Cumulative symptomatic cases at day 250 |  | Date of peak symptomatic prevalence |  |
| --- | --- | --- | --- | --- | --- | --- |
|  | (# cases) | Reduction from baseline (#/% cases) | (# cases) | Reduction from baseline (#/% cases) | (Date of epidemic) | Delay from baseline (# days) |
| Baseline, no intervention | 75,809 |  | 228,030 |  | 131 |  |
| STQ- Isolate tested-positive, symptomatic, and household: |  |  |  |  |  |  |
| 10K tests | 24,734 | 51,075 / 67% | 135,925 | 92,105 / 40% | 181 | 50 |
| 50K tests | 17,683 | 58,126 / 77% | 105,643 | 122,387 / 54% | 205 | 74 |
| 100K tests | 9,459 | 66,351 / 88% | 53,569 | 174,461 / 77% | 239 | 108 |
| 200K tests | 342 | 75,467 / 99.6% | 2,966 | 225,064 / 99% | 241 | 110 |

<sup>1</sup> All values represent the average of 10 model runs, each starting with a different random seed.
<sup>2</sup> All STQ scenarios used simple random sampling, tests every seven days.

**Table S3.** Efficacy of *STQ* interventions, by frequency of testing. Scenarios here are the same as those presented in Figure 3 in the main article.

|  | Symptomatic prevalence at peak |  | Cumulative symptomatic cases at day 250 |  | Date of peak symptomatic prevalence |  |
| --- | --- | --- | --- | --- | --- | --- |
|  | (# cases) | Reduction from baseline (#/% cases) | (# cases) | Reduction from baseline (#/% cases) | (Date of epidemic) | Delay from baseline (# days) |
| Baseline, no intervention | 75,809 |  | 228,030 |  | 131 |  |
| STQ- Isolate tested-positive, symptomatic, and household |  |  |  |  |  |  |
| 10K tests, daily frequency | 16,482 | 59,327 / 78% | 100,467 | 127,564 / 56% | 211 | 80 |
| 50K tests, 7 day frequency | 17,683 | 58,126 / 77% | 105,643 | 122,387 / 54% | 205 | 74 |
| 100K tests, 14 day frequency | 16,599 | 59,211 / 78% | 96,164 | 131,866 / 58% | 214 | 83 |
| 200K tests, 28 day frequency | 16,743 | 59,066 / 78% | 95,603 | 132,427 / 58% | 211 | 80 |

<sup>1</sup> All values represent the average of 10 model runs, each starting with a different random seed.
<sup>2</sup> All STQ scenarios used simple random sampling.

**Table S4.** Efficacy of STQ interventions, by sampling strategy. Scenarios here are the same as those presented in Figure 4 in the main article.

|  | Symptomatic prevalence at peak |  | Cumulative symptomatic cases at day 250 |  | Date of peak symptomatic prevalence |  |
| --- | --- | --- | --- | --- | --- | --- |
|  | (# cases) | Reduction from baseline (#/% cases) | (# cases) | Reduction from baseline (#/% cases) | (Date of epidemic) | Delay from baseline (# days) |
| Baseline, no intervention | 75,809 |  | 228,030 |  | 131 |  |
| STQ- Isolate tested-positive, symptomatic, and household members |  |  |  |  |  |  |
| Simple random sampling | 17,683 | 58,126 / 77% | 105,643 | 122,387 / 54% | 205 | 74 |
| Cluster sampling on communities | 17,394 | 58,415 / 77% | 107,084 | 120,947 / 53% | 190 | 58 |
| Cluster sampling on schools | 9,745 | 66,064 / 87% | 52,201 | 175,829 / 77% | 241 | 110 |
| Pooled sampling, 2-person pools | 9,900 | 65,909 / 87% | 57,698 | 170,332 / 75% | 236 | 105 |
| Pooled sampling, 5-person pools | 129 | 75,681 / 99.8% | 871 | 227,159 / 99.6% | 78 | -54 |

\* All values represent the average of 10 model runs, each starting with a different random seed.

## Footnotes

1 Hereinafter, for brevity we use the term “asymptomatic” to denote those who are infected with SARS-COV-2, are infectious, but are not currently experiencing noticeable symptoms, either because they are pre-symptomatic or asymptomatic.

2 Available at: https://www.cdc.gov/coronavirus/2019-ncov/covid-data/covidview/index.html, accessed September 9, 2020.

3 Available at: https://www.cdc.gov/coronavirus/2019-ncov/covid-data/covidview/index.html, accessed September 10, 2020.

## References

1. O. Byambasuren et al., Estimating the Extent of True Asymptomatic COVID-19 and Its Potential for Community Transmission: Systematic Review and Meta-Analysis Available at SSRN: https://ssrn.com/abstract=3586675 or http://dx.doi.org/10.2139/ssrn.3586675, (2020).

2. Centers for Disease Control and Prevention, “COVID-19 Pandemic Planning Scenarios,” (Centers for Disease Control, https://www.cdc.gov/coronavirus/2019-ncov/hcp/planning-scenarios.html, 2020).

3. M. Chinazzi et al., The effect of travel restrictions on the spread of the 2019 novel coronavirus (COVID-19) outbreak. Science, eaba9757 (2020).

4. C. Heneghan, J. Brassey, T. Jefferson, “COVID-19: What proportion are asymptomatic?,” (Center for Evidence-Based Medicine, https://www.cebm.net/covid-19/covid-19-what-proportion-are-asymptomatic/, 2020).

5. S. G. Krantz, A. S. R. S. Rao, Level of under-reporting including under-diagnosis before the first peak of COVID-19 in various countries: Preliminary Retrospective Results Based on Wavelets and Deterministic Modeling. Infection Control & Hospital Epidemiology, 1–8 (2020).

6. R. Li et al., Substantial undocumented infection facilitates the rapid dissemination of novel coronavirus (SARS-CoV2). Science, eabb3221 (2020).

7. M. Morris, “Estimating total COVID-19 infected persons in King County, WA,” (http://statnet.org/CovidBackCalc/KCReport.html, 2020).

8. M. Park, A. R. Cook, J. T. Lim, Y. Sun, B. L. Dickens, A Systematic Review of COVID-19 Epidemiology Based on Current Evidence. Journal of Clinical Medicine 9, 967 (2020).

9. J. Hellewell et al., Feasibility of controlling COVID-19 outbreaks by isolation of cases and contacts. The Lancet Global Health 8, e488–e496 (2020).

10. M. J. Keeling, T. D. Hollingsworth, J. M. Read, Efficacy of contact tracing for the containment of the 2019 novel coronavirus (COVID-19). Journal of Epidemiology and Community Health doi: 10.1136/jech-2020-214051, (2020).

11. J. M. Epstein, Modelling to contain pandemics. Nature 460, 687–687 (2009).

12. R. Hinch et al., OpenABM-Covid19 - an agent-based model for non-pharmaceutical interventions against COVID-19 including contact tracing. medRxiv, 2020.2009.2016.20195925 (2020).

13. N. Hoertel et al., A stochastic agent-based model of the SARS-CoV-2 epidemic in France. Nature Medicine 26, 1417–1421 (2020).

14. N. Mahdizadeh Gharakhanlou, N. Hooshangi, Spatio-temporal simulation of the novel coronavirus (COVID-19) outbreak using the agent-based modeling approach (case study: Urmia, Iran). Informatics in Medicine Unlocked 20, 100403 (2020).

15. L. Perez, S. Dragicevic, An agent-based approach for modeling dynamics of contagious disease spread. International Journal of Health Geographics 8, 50 (2009).

16. P. C. L. Silva et al., COVID-ABS: An agent-based model of COVID-19 epidemic to simulate health and economic effects of social distancing interventions. Chaos, Solitons & Fractals 139, 110088 (2020).

17. S. Venkatramanan et al., Using data-driven agent-based models for forecasting emerging infectious diseases. Epidemics 22, 43–49 (2018).

18. E. Karimi, K. Schmitt, A. Akgunduz, Effect of individual protective behaviors on influenza transmission: an agent-based model. Health Care Management Science 18, 318–333 (2015).

19. S. Kumar, J. J. Grefenstette, D. Galloway, S. M. Albert, D. S. Burke, Policies to Reduce Influenza in the Workplace: Impact Assessments Using an Agent-Based Model. American Journal of Public Health 103, 1406–1411 (2013).

20. S. Merler et al., Spatiotemporal spread of the 2014 outbreak of Ebola virus disease in Liberia and the effectiveness of non-pharmaceutical interventions: a computational modelling analysis. The Lancet Infectious Diseases 15, 204–211 (2015).

21. A. Aleta et al., Modelling the impact of testing, contact tracing and household quarantine on second waves of COVID-19. Nature Human Behaviour 4, 964–971 (2020).

22. N. Hoertel et al., Facing the COVID-19 epidemic in NYC: a stochastic agent-based model of various intervention strategies. medRxiv : the preprint server for health sciences, 2020.2004.2023.20076885 (2020).

23. D. L. Chao, A. P. Oron, D. Srikrishna, M. Famulare, “Modeling layered non-pharmaceutical interventions against SARS-COV-2 in the United States with CORVID,” (Preprint available at https://www.medrxiv.org/content/10.1101/2020.04.08.20058487v1, 2020).

24. Y. Alimohamadi, M. Taghdir, M. Sepandi, Estimate of the Basic Reproduction Number for COVID- 19: A Systematic Review and Meta-analysis. Journal of Preventive Medicine & Public Health 53, 151–157 (2020).

25. M. Irvine et al., Modeling COVID-19 and Its Impacts on U.S. Immigration and Customs Enforcement (ICE) Detention Facilities, 2020. Journal of Urban Health 97, 439–447 (2020).

26. M. Lonergan, J. D. Chalmers, Estimates of the ongoing need for social distancing and control measures post-”lockdown” from trajectories of COVID-19 cases and mortality. European Respiratory Journal, 2001483 (2020).

27. S. M. Moghadas et al., Projecting hospital utilization during the COVID-19 outbreaks in the United States. Proceedings of the National Academy of Sciences 117, 9122–9126 (2020).

## SI References

1. Y. Alimohamadi, M. Taghdir, M. Sepandi, Estimate of the Basic Reproduction Number for COVID- 19: A Systematic Review and Meta-analysis. Journal of Preventive Medicine & Public Health 53, 151–157 (2020).

3. M. Lonergan, J. D. Chalmers, Estimates of the ongoing need for social distancing and control measures post-”lockdown” from trajectories of COVID-19 cases and mortality. European Respiratory Journal, 2001483 (2020).

4. M. Irvine et al., Modeling COVID-19 and Its Impacts on U.S. Immigration and Customs Enforcement (ICE) Detention Facilities, 2020. Journal of Urban Health 97, 439–447 (2020).

5. S. M. Moghadas et al., Projecting hospital utilization during the COVID-19 outbreaks in the United States. Proceedings of the National Academy of Sciences 117, 9122–9126 (2020).

